# Foundation Model Artificial Intelligence for Electrocardiogram Detection of Rheumatic Mitral Stenosis

**DOI:** 10.64898/2026.10.12.26365300

**Authors:** Raghav Sriram, Ivan Nenadic, Zak Loring, Marat Fudim, Anthony J. Mazella, Venkatesh L. Murthy, Anil K. Gehi

**Author notes:** To whom correspondence should be sent. **Corresponding Author:** Dr. Anil Gehi, Electronic address.

## Abstract

**Background:** Rheumatic mitral stenosis (RMS) disproportionately affects low-resource settings where echocardiography is limited. We evaluated foundation model (FM) feasibility for RMS detection using limited, indirectly labeled ECGs.

**Methods:** An ECG-FM was fine-tuned using 804 RMS-positive ECGs per fold and externally validated across clinical and wearable lead configurations.

**Results:** Internal AUROCs were 0.89, 0.87, and 0.87, for 12-Lead, 6-Lead, and 1-Lead respectively. External AUROCs were 0.82, 0.80, and 0.77. Performance remained comparable across demographic subgroups.

**Conclusions:** FM fine-tuning enables RMS detection using limited ICD-derived labels, supporting further evaluation of reduced-lead ECG screening and echocardiography triage in resource-constrained settings.

## Main Text

Rheumatic Mitral Stenosis (RMS) is the most common lesion in chronic Rheumatic Heart Disease (RHD) ^1^ and is widely prevalent in Low or Middle Income Countries (LMICs). Limited echocardiogram access in LMICs often results in delayed RMS diagnosis and adverse outcomes. ECG-AI could increase RMS diagnostic access in LMICs. Prior RHD ECG-AI papers oversample ECGs from a limited number of positive patients ^2,3^, do not focus on RMS ^2,3^, are not externally-validated ^2,3^, and do not evaluate reduced lead formats ^2,3^. Limited RMS prevalence in Western nations hinders ECG-AI development utilizing traditional methods ^2^. Advances in ECG-AI show foundation model (FM) fine-tuning improves rare-disease ECG detection ^2^ and FM’s pretrained with lead-masking support generalization across lead formats ^4^. Given limited access to large, annotated ECG datasets in RHD-endemic LMICs, our approach demonstrates the feasibility of developing generalizable RMS classifiers for clinical and wearable applications using relatively few labeled ECGs and indirect disease labels derived from diagnostic codes from United States cohorts.

From the Harvard-Emory ECG Database ^5^, we selected training ECGs from Massachusetts General Hospital (MGH) and externally validated on ECGs from Emory University Hospital (EUH). RMS patients were identified with ICD-9 code 394.0 or ICD-10 code I05.0. We selected each patient’s closest ECG preceding ICD diagnosis. ECGs more than 2 years prior to diagnosis were excluded. Mean ECG time for MGH was 4.57 ± 5.39 months and 5.74 ± 7.04 for EUH. Control ECGs were selected from unique patients without any coded RHD or RMS. ECGs were z-score normalized across leads. Limb lead (6L) and Lead 1 (1L) configurations mimicking commercially available devices were created by duplicating the 12-Lead (12L) folds and filling unavailable leads with zero. Five-fold cross-validation was performed with a balanced training cohort. Each fold’s training set contained a random sample of 804 positive ECGs and 804 negative ECGs. Validation and internal test sets contained 10,000 and 10,100 ECGs respectively with each having 100 positive ECGs to reflect a prevalence of 1% found in some endemic nations. External validation also reflected this prevalence with 529 positive ECGs and 52,349 negative ECGs. Both cohorts were approximately 52% female. Mean age in MGH was 54.65 ± 20.80, and mean age in EUH was 58.32 ± 16.72. The MGH cohort was 71% white patients, with the most prevalent defined minority group being black patients at 6%. The EUH cohort was 55% white patients, 32% black patients, and 13% from other racial groups.

We utilized ECG-FM ^4^, a transformer-based FM with 90,374,017 parameters pretrained with generative masking, contrastive multi-segment coding, and random lead masking. The FM was fine-tuned using an NVIDIA RTX 2000 GPU with an Adam optimizer at a fixed learning rate of 1e-06, a training batch size of 32, and binary cross-entropy loss. Training was set for 140 epochs and stopped if validation loss did not improve for 7 epochs.

Internally AUROCs were 0.89 [0.88, 0.90], 0.87 [0.86, 0.88], and 0.87 [0.86, 0.88] with AUPRCs of 0.08 [0.07, 0.09], 0.07 [0.06, 0.08], and 0.08 [0.07, 0.09] for 12L, 6L, and 1L respectively. EUH external AUROCs were 0.82 [0.81, 0.83], 0.80 [0.78, 0.82], and 0.77 [0.75, 0.79] with AUPRCs of 0.05 [0.04, 0.06], 0.04 [0.03, 0.05], and 0.04 [0.03, 0.05] for 12L, 6L, and 1L respectively. Screening results from a fixed threshold where the models achieved a validation sensitivity of 80% across lead-configurations and a saliency map of importance attribution on the MGH test set are shown in the figure.

**Figure 1.**
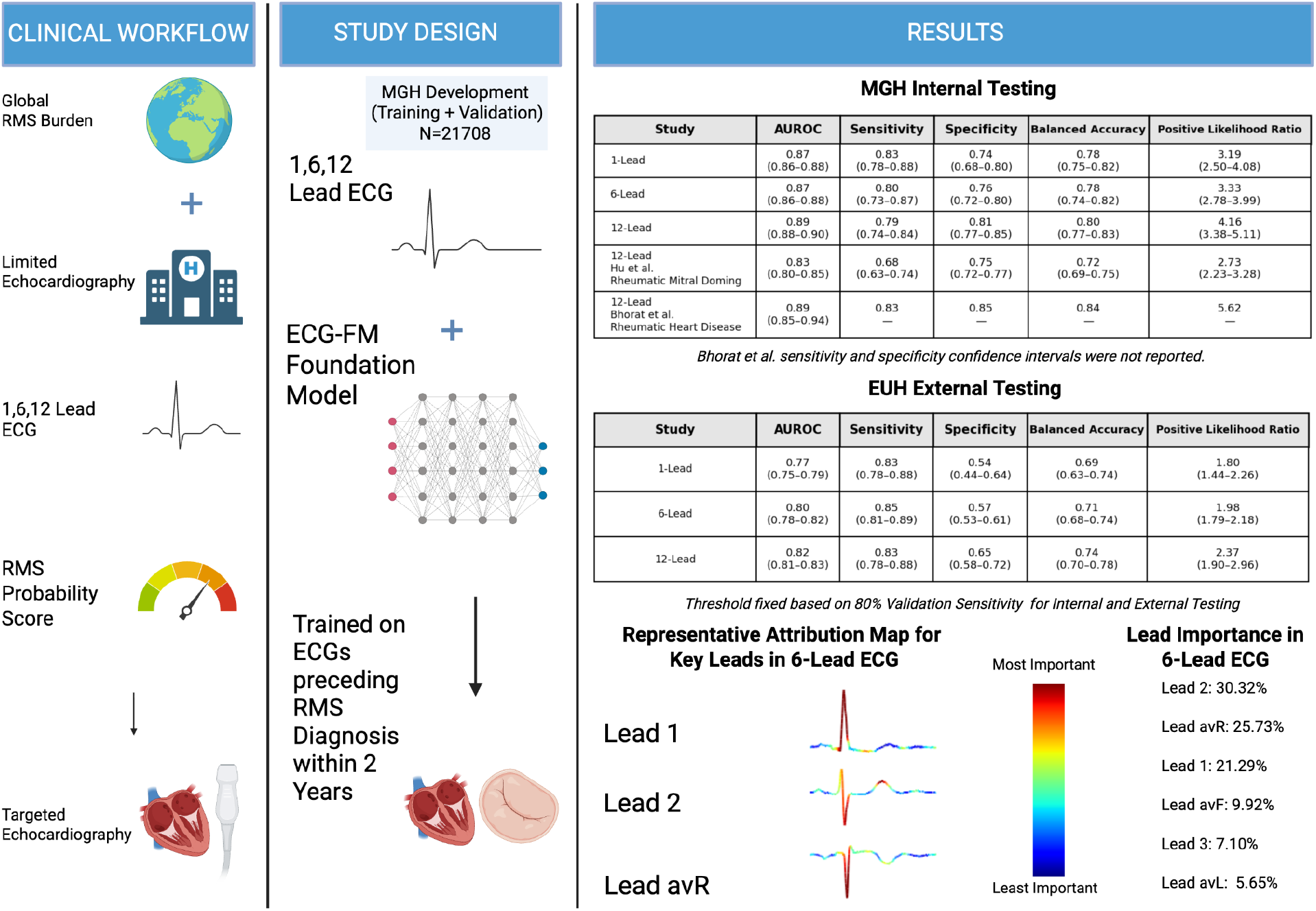
Study design for evaluating the performance of clinical and wearable ECG-RMS. Created in BioRender. Sriram, R. (2026) https://BioRender.com/0laqdnf

Performance was comparable across lead-configuration, sex, above and below age 60, and white, black, and other race patients. Preservation of sensitivity across lead-formats supports the feasibility of limited-lead ECG screening on devices like KardiaMobile 6L. However, reduced external specificity indicates a high-sensitivity strategy could generate substantial false-positives, indicating this approach is best viewed as an echocardiography triage tool rather than a stand-alone diagnostic test.

Limitations to this study included ICD-based RMS labels due to lack of publicly available echocardiogram-confirmed RMS ECGs. Our models should be interpreted as detecting ICD-coded RMS rather than echocardiography adjudicated mitral stenosis. Additionally, control patients were defined by absence of RHD codes, which may not exclude undiagnosed or miscoded disease. Although reduced-lead performance was encouraging, specificity declined externally, and prospective evaluation on low-cost edge devices is needed to determine computational efficiency, downstream echocardiography burden, and clinical utility. Exploratory attribution mapping localizing on the QRS should not be interpreted as definitively causal features. Finally, validation in LMICs is essential, as disease spectrum, comorbidities, ECG acquisition conditions, and pretest probability may differ from US cohorts.

Our findings with standard and reduced-lead ECGs support further evaluation of ECG-RMS for screening and echocardiography triage in resource-constrained settings with RHD burden.

## Data Availability

All data produced are available online at the Harvard-Emory ECG Database: https://bdsp.io/content/heedb/5.0/

## References

1. Pandian NG, Kim JK, Arias-Godinez JA, et al, Recommendations for the Use of Echocardiography in the Evaluation of Rheumatic Heart Disease: A Report from the American Society of Echocardiography. J Am Soc Echocardiogr. 2023 Jan;36(1):3–28. doi: 10.1016/j.echo.2022.10.009. Epub 2022 Nov 23. Erratum in: J Am Soc Echocardiogr. 2023 Apr;36(4):445. doi: 10.1016/j.echo.2023.01.016. PMID: 36428195.

2. Stephanie M Hu, Joshua P Barrios, Geoffrey H Tison, A deep foundation model for electrocardiogram interpretation: enabling rare disease detection through transfer learning, European Heart Journal - Digital Health, Volume 6, Issue 4, July 2025, Pages 619–623, 10.1093/ehjdh/ztaf051

3. Raeesa Bhorat, David M Harmon, Kan Liu, et al, Artificial Intelligence Enhanced Electrocardiogram for the Detection of Rheumatic Heart Disease, European Heart Journal - Digital Health, 2026;, ztag056, 10.1093/ehjdh/ztag056

4. Kaden McKeen, Sameer Masood, Augustin Toma, Barry Rubin, Bo Wang, ECG-FM: an open electrocardiogram foundation model, JAMIA Open, Volume 8, Issue 5, October 2025, ooaf122, 10.1093/jamiaopen/ooaf122

5. Koscova, Z., Moura Junior, V., Reyna, M., et al. (2026). Harvard-Emory ECG Database (version 5.0). Brain Data Science Platform. 10.60508/rv6h-7d10.

